# Biopsychosocial and Demographic Predictors of Functional Brain Network Specialization and Segregation Across the Adult Lifespan

**DOI:** 10.1101/2025.11.20.25340014

**Authors:** A. Shankar, M. Phansikar, N. McPherson, A.X. Gorka, B.M. Way, R. Betzel, R.S. Prakash

## Abstract

The properties of functional brain networks are an important determinant of cognitive function in aging and dementia. Despite this, few studies have comprehensively examined demographic and biopsychosocial predictors of functional brain networks, and none have attempted to do so across the adult lifespan while accounting for collinearity among these predictors. The current study used data from 525 individuals between the ages of 35 and 100 years from the Human Connectome Project 2.0 Lifespan Release, which includes task-based functional neuroimaging, physical and emotional health, and demographic information. Two functional brain network properties previously identified as moderators of cognitive decline—entropy and modularity—were used as outcome metrics in four elastic net regression models that identified and ranked predictors of these metrics as well as their age-interaction terms. We identified biological sex, sleep duration, instrumental support, visual acuity, education, social isolation, diastolic blood pressure, and vigorous physical activity as the strongest and most consistent predictors of entropy and modularity beyond age. Importantly, these predictors differed from ranked correlational results, suggesting many predictors share large amounts of overlapping statistical variance. Additionally, we found that biological sex exhibited a significant moderation effect such that males demonstrated greater age-related decreases to network resilience with increasing age compared to females. In the current study, we ranked biopsychosocial health determinants of network properties in an adult lifespan sample. Given previous research implicating modularity and entropy as possible measures of cognitive reserve, these results may inform our understanding of resilience to cognitive decline in aging.

## 1. Introduction

An estimated 153 million people worldwide will be living with dementia by the year 2050 (Davis et al., 2008; Maillet & Rajah, 2014). Epidemiological studies and aggregated randomized controlled trials have suggested that at least 45% of dementia risk is preventable by optimizing wellness in several key domains across the lifespan, including cardiovascular, physical, and mental health (Barnes & Yaffe, 2011; Livingston et al., 2024), making efforts to understand the mechanisms of how modifying risk factors may reduce dementia incidence an important priority. Leading cognitive aging models emphasize the complex and interdependent relationship between biopsychosocial risk factors and dementia-related mechanisms, such as vascular disease, accumulation of key proteinopathies, and inflammation (Josefsson et al., 2012; Livingston et al., 2024; Reuter-Lorenz & Park, 2014; Tangestani Fard & Stough, 2019). Additionally, these models have suggested that functional brain networks are likely both a pathway through which these pathological changes result in cognitive decline, as well as an important mechanism of conferring resilience, referred to as cognitive reserve (Livingston et al., 2024; Reuter-Lorenz & Park, 2014).

For example, previous literature and our recent work has identified age-related changes in two key graph theory metrics—modularity, a measure of network segregation, and entropy, a measure of nodal specialization. Both metrics strongly covaried with age (Puxeddu et al., 2020; Shankar et al., 2024; Song et al., 2014) and moderated cognitive performance in older adults such that individuals with greater network segregation and specialization demonstrated better cognitive performance (Chan et al., 2021; Shankar et al., 2024). In children and adolescents, socioeconomic status, education, and environmental richness predict trajectories and endpoints of network segregation (Tooley et al., 2021). In older adults, educational attainment has similarly been implicated as a protective factor for network resilience and cognitive decline (Chan et al., 2021; Kim et al., 2021). Additionally, research has suggested that functional brain network changes in aging are also a mechanism of cognitive decline (Gamboa et al., 2014; Lebedev et al., 2014; Nashiro et al., 2017). Thus, certain biopsychosocial factors may reduce disease pathology and its consequences to functional brain networks, and other factors may increase resilience of functional brain networks in such a way that reduces the observable impact of that pathology. Despite this important dual function of brain networks for cognitive performance and quality of life, most research has focused on identifying risk and protective factors of cognition and dementia, and relatively little work has examined risk and protective factors of brain network function directly. This work is important both for personalized medicine approaches and public health efforts to preventing cognitive decline in aging.

Recent work has begun to examine additional correlates of graph theory metrics to better identify the specific demographic and lifestyle factors that may influence network development and maintenance. These studies have identified some common correlates of aging, such as poor cardiovascular health, metabolic health, and sleep deprivation with decreased network segregation (Ben Simon et al., 2017; Kong et al., 2020; Manza et al., 2020; Rashid et al., 2021). Other factors, such as emotional well-being, stress, and psychiatric symptoms of anxiety and depression have been linked to cognitive decline (Botto et al., 2022) but less well studied in relation to functional brain networks (Gong & He, 2015a; R. Wang et al., 2022; Yang et al., 2019; Ye et al., 2015; Yun & Kim, 2021; Zhang et al., 2020). Taken together, this evidence suggests that many facets of demographics and biopsychosocial health may influence the development and maintenance of functional brain networks. However, there is currently significant gaps in the literature comprehensively examining biological, psychological, and social factors simultaneously, understanding how these highly collinear factors influence brain networks differentially, and how these relationships may change across the lifespan. We propose a need to identify determinants of network organization in a data-driven manner that acknowledges the collinearity between these variables and their interactions with age across the adult lifespan. In the current study, we used elastic-net regression, a machine learning method that allows for collinearity among predictor variables, as a feature selection tool to identify and rank modifiable and demographic predictors of two functional network properties, whole-brain modularity and entropy, that either uniquely affect these network properties or potentially moderate our previously reported age-related changes to network integrity. We also included models with age-interaction terms to determine which variables may demonstrate age-related changes in their relationship to these network properties.

## 2. Methods

### 2.1 Participants and Procedures

The initial sample for this study consisted of cross-sectional data from 725 healthy individuals ages 34-100+ years old (55.7% female) from the Lifespan Human Connectome Project Aging (HCP-A) 2.0 Release in 2021. For a description of the dataset, see the Supplemental Methods section and the study protocols (Bookheimer et al., 2019; Harms et al., 2018). From this dataset, we included a total of 28 predictor variables (Table 1), described in detail in the Supplemental Methods, including demographic data, blood draw measures, physical exam data, and self-report measures.

**Table 1.** Descriptive Statistics of Model Predictor Variables.

| Predictor | Total (n = 525) | Female (n = 295) | Male (n = 230) |
| --- | --- | --- | --- |
| Age (years) | 59.67 (15.13) | 59.87 (15.38) | 59.41 (14.83) |
| Education (years) | 15.81 (2.07) | 15.66 (2.12) | 16 (1.98) |
| Depression (raw score) | 9.01 (2.65) | 9.01 (2.65) | 9 (2.66) |
| Perceived Stress (raw score) | 22.3 (5.66) | 22.2 (5.56) | 22.44 (5.79) |
| Affective Anxiety (raw score) | 9.08 (2.34) | 9.05 (2.35) | 9.12 (2.33) |
| Somatic Anxiety (raw score) | 7.95 (2.26) | 7.99 (2.12) | 7.89 (2.43) |
| Instrumental Support (raw score) | 31.26 (8.62) | 30.82 (8.48) | 31.83 (8.78) |
| Social Isolation (raw score) | 9.19 (3.73) | 8.95 (3.53) | 9.5 (3.96) |
| Life Satisfaction (raw score) | 22.8 (9.98) | 23.26 (10.89) | 22.21 (8.66) |
| Meaning and Purpose (raw score) | 27.76 (14.33) | 28.39 (14.41) | 26.95 (14.21) |
| Pain Intensity (raw score) | 1.88 (1.98) | 1.94 (2.04) | 1.8 (1.89) |
| Pain Interference (raw score) | 9.31 (3.26) | 9.3 (3.46) | 9.33 (2.99) |
| Auditory Acuity (raw score) | 20.81 (5.55) | 21.48 (5.33) | 19.95 (5.72) |
| Visual Acuity (raw score) | 84.16 (6.88) | 83.83 (6.93) | 84.59 (6.79) |
| Walking (days/week) | 5.08 (2.34) | 5.16 (2.33) | 4.98 (2.35) |
| Moderate Physical Activity (days/week) | 2.61 (2.35) | 2.66 (2.36) | 2.54 (2.34) |
| Vigorous Physical Activity (days/week) | 2.04 (2.16) | 1.73 (2.03) | 2.44 (2.26) |
| Sleep Duration (hours/night) | 6.81 (1.08) | 6.89 (1.09) | 6.71 (1.05) |
| Sleep Efficiency (%) | 88.92 (12.43) | 88.72 (12.67) | 89.19 (12.14) |
| Triglycerides (mg/dL) | 108.01 (67.68) | 100.43 (62.04) | 117.73 (73.29) |
| Cholesterol (mg/dL) | 198.19 (36.06) | 202.58 (33.52) | 192.57 (38.43) |
| hsCRP (mg/L) | 2.42 (4.49) | 2.64 (5.22) | 2.13 (3.3) |
| A1crs (%) | 5.37 (0.53) | 5.33 (0.51) | 5.41 (0.56) |
| Systolic Blood Pressure (mm Hg) | 129.21 (16.55) | 127.39 (16.8) | 131.54 (15.97) |
| Diastolic Blood Pressure (mm Hg) | 79.39 (10.55) | 77.42 (10.07) | 81.93 (10.63) |
| BMI (kg/m<sup>2</sup>) | 26.99 (4.76) | 26.76 (5.22) | 27.28 (4.1) |
| Emotional Support (raw score) | 33.81 (5.79) | 34.37 (5.56) | 33.1 (6.01) |
| Pulse Pressure (mm Hg) | 49.81 (13.38) | 49.97 (13.98) | 49.61 (12.58) |
| Entropy | 0.78 (0.03) | 0.77 (0.03) | 0.78 (0.03) |
| Modularity | 0.5 (0.04) | 0.5 (0.03) | 0.49 (0.04) |
| Total Brain Volume (mm<sup>3</sup>) | 1,529,974.85<br>(191,638.81) | 1,435,737.38<br>(161,414.43) | 1,650,844.66<br>(156,435.11) |

Note: hsCRP = high sensitivity C-reactive Protein. BMI = body mass index.

### 2.2 Neuroimaging Outcome Variables

The fMRI data was minimally preprocessed by the HCP study coordinators (see the processing protocol (Glasser et al., 2013) for a detailed pipeline) using Freesurfer and a custom ICA-Fix pipeline developed specifically for and validated on the HCP datasets. This process resulted in cortical data in grayordinate space and subcortical data in volumetric space. For the current study, we additionally removed global signal using MATLAB by averaging across all cortical grayordinates and regressing this signal from the data. This data was parcellated using the recommended atlas for the dataset: the Glasser Atlas for the cortical data and Cole Atlas for subcortical data, consisting of 360 cortical regions and 19 subcortical regions.

For a detailed description of modularity and entropy calculations for the current project, see the methods in our previous work (Shankar et al., 2024). Briefly, we used the Louvain algorithm for community detection in the Brain Connectivity Toolbox (Rubinov & Sporns, 2010) to calculate modularity and an edge-connectivity approach (Faskowitz et al., 2020) to calculate nodal entropy. Brain network modules consist of groupings of brain regions, or nodes, that are densely interconnected while being much less connected to other modules; therefore modularity is a measure of the degree of inter-versus intra-network connectivity among all modules in a network (Sporns & Betzel, 2016). For entropy, we used an edge-centric framework to cluster edges based on their temporal similarity during the scanning period, which can be used to reveal an overlapping community structure (Faskowitz et al., 2020; Jo et al., 2021). The distribution of participation for each node in each community can then be used to calculate entropy as the fraction of each node’s edges that belong to a community multiplied by the log of that fraction summed over all communities. Entropy can therefore be conceptualized as a measure of nodal specialization across these communities. For the primary analyses, we used whole-brain modularity and whole-brain averaged entropy as our outcome metrics.

To control for the potentially confounding role of in-scanner head motion, we regressed mean framewise displacement values from entropy and modularity calculations. Previous work using fMRI data in machine learning analyses has recommended that nuisance variables be regressed from outcome variables prior to model fitting as the most stringent method of controlling for noise, but also recommend reporting models with no nuisance regression (Pervaiz et al., 2020). Here, we report the former as our primary analyses and the latter as supplementary.

### 2.3 Elastic Net Regression

All biopsychosocial variables were examined for collinearity, near zero variance, and outliers. Collinearity was examined using a correlation matrix to examine the direction and strength of the raw relationships between the predictor variables and functional connectivity metrics. We examined the near zero variance of all predictors, screening for those that are likely to contribute little predictive value due to having few unique values relative to the number of observations. Given that machine learning algorithms may be biased toward overfitting by extreme outliers (Wujek et al., 2016), all biopsychosocial predictor variables were winsorized such that values greater than 3 standard deviations from the mean were replaced with the closest non-outlier value.

Prior to running the regression, 30% of the data was withheld as a validation sample, selected randomly and stratified by age. The remaining 70% of the data was used to derive an optimal fit model by iteratively splitting the data further into training and testing sets and identifying model features and optimal values for the model parameters of alpha and lambda using the “caret” package (Kuhn, 2008) and “glmnet” package (Friedman et al., 2010) in R Studio version 2022.07.2. Alpha, bounded from 0 to 1, determines the sparsity of predictor inclusion for deriving the best fit model; models with an alpha of 1 are equivalent to lasso regression, and models with an alpha of 0 are equivalent to ridge regression. Lambda determines the degree of regularization applied to the model to reduce overfitting (Zou & Hastie, 2005).

### 2.4 Model training and testing

The elastic net models used a repeated 10-fold cross-validation procedure that randomly split the data into 10 subgroups and iteratively held one group as a testing set while training on the remaining data, until all ten groups served as the test set. For each k-fold, data was centered and scaled. Model hyperparameters were tested by searching a grid of 100 lambda values of 10^-5^ to 1 and 21 alpha values between 0 and 1. This process was repeated over 100 repetitions with each repetition shuffling the data into new groups. For each k-fold, we tested 2100 models, repeated 100 times. From these iterations, the alpha and lambda combination with the lowest average root mean squared error were selected as the hyperparameters for the best model. These hyperparameters were then applied to the training sample of the dataset to determine the selected features and their weights. The accuracy of this model on the 70% training set was measured as the r-squared of network metrics values as predicted by this model and their true values as well as the mean squared error (MSE).

### 2.5 Evaluation of model feature consistency

To determine the consistency of features selected across alpha and lambda combinations, we examined the selected coefficients in the best-performing 10% of models, as measured by having the lowest root mean squared error. For each feature, we calculated the percentage of iterations it was selected for in these 10% of models.

### 2.6 Model generalization to validation data

Following model derivation as outlined above on 70% of the data, the best fit model was applied to the held-out validation sample to estimate the generalizability of the model’s predictors, weights, and hyperparameters. Model accuracy was measured as the r-squared and the mean squared error of network entropy and modularity as predicted by the derived model on this unseen data compared to the true values of entropy and modularity.

### 2.7 Regional Entropy Values of Selected Variables

To examine the relationship between biopsychosocial variables and regional brain function, we correlated regional entropy values at the level of 379 individual brain nodes with the values for the final model-selected variables. We used false discovery rate correction to identify the individual brain regions with the strongest relationship to the predictor variables.

## 3. Results

### 3.1 Sample Demographics and Descriptive Statistics

The final dataset consisted of 525 adults who had complete data for the neuroimaging outcome variables and the biopsychosocial predictors of interest. Demographically, these participants had an average age of 59.67 years (*SD* = 15.13), 15.81 years of education (*SD* = 2.07), and 56.19% of the sample identified as female. Additional descriptive statistics can be found in Table 1. We observed collinearity among the biopsychosocial predictors, particularly within cardiovascular health variables and within emotional health variables (Figure 1a). We also examined the zero-order associations between our predictors and functional connectivity metrics, finding that in addition to age, auditory acuity, visual acuity, and blood pressure had the strongest relationships to both entropy and modularity (Figure 1b). None of our metrics demonstrated a near zero variance, and therefore all were included as model predictors.

**Figure 1.**
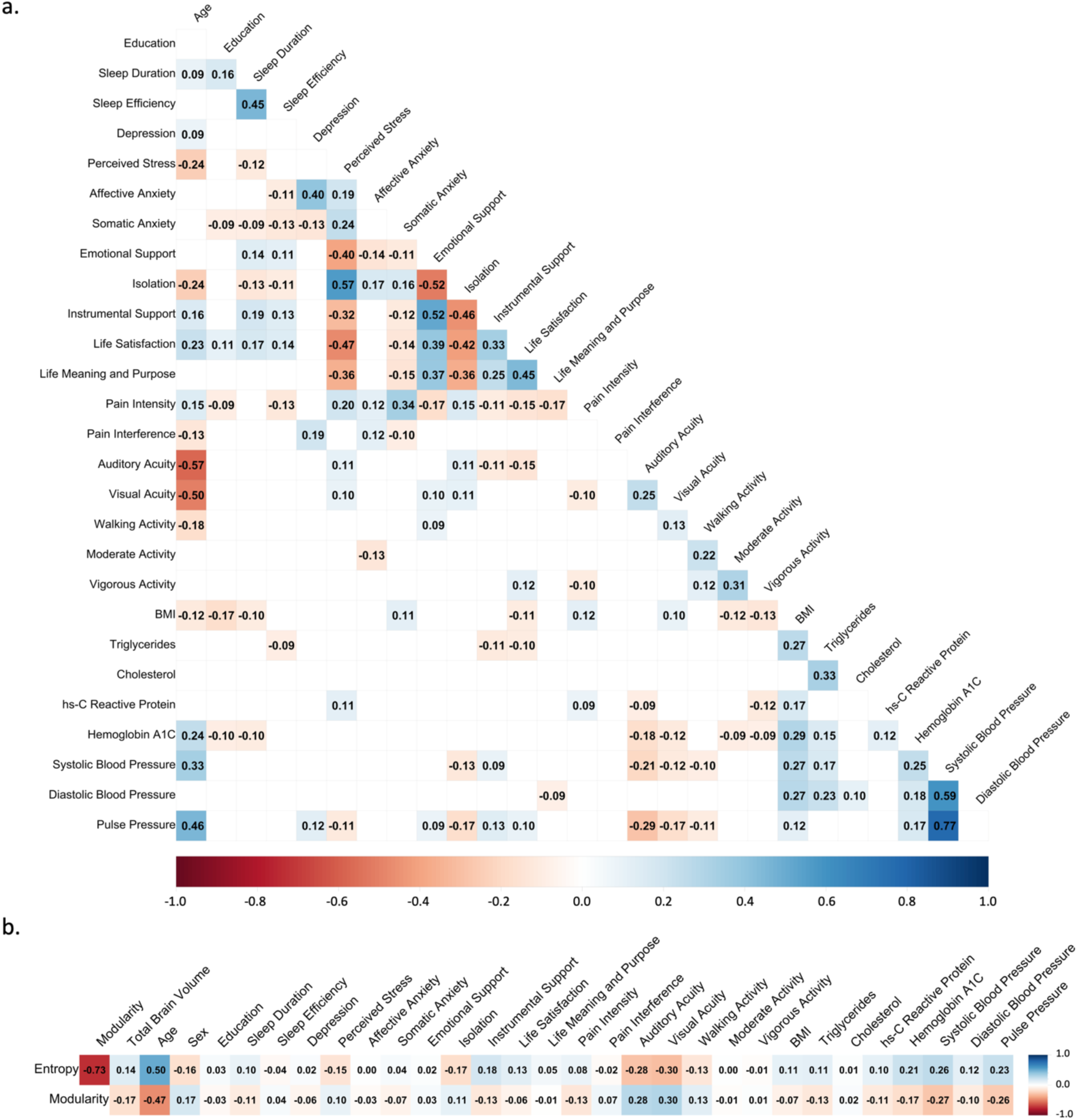
Correlations among predictor variables and outcome variables. a) Correlation plot of all biopsychosocial variables. Only correlations that are significant with p < 0.05 are shown. b) Correlation plot between predictor variables and modularity and entropy. BMI = Body Mass Index.

### 3.2 Simple Model Results

We examined the selected predictors for the top 10% of all simple models and report the percent of these top models that each variable was selected for (Figure 2a & Figure 2b). For entropy, the best fitting model from the derivation procedure identified nine final predictor variables (α = 0.6, λ = 0.0021). These variables, ranked in order of importance to model predictive power, were age, sex, sleep duration, instrumental support, visual acuity, education, isolation, diastolic blood pressure, and vigorous physical activity (Figure 3a). This model’s performance when applied to the 369 individuals in the training set yielded an r-squared of 0.28 (R= 0.53) and yielded a similar r-squared of 0.25 (R=0.50, MSE = .00054) when applied to the 156 individuals in the held-out validation set. For modularity, the best fitting model identified eight final predictor variables (α = 1, λ = 0.0024) (Figure 3b), ranked as: age, sex, sleep duration, visual acuity, education, instrumental support, affective anxiety, and diastolic blood pressure. Of note, the alpha value of this model of 1 suggests the model performed best as a lasso model. The model when applied to the 369 individuals in the training set yielded a slightly lower r-squared of 0.22 (R= 0.47) and an r-squared of 0.25 (R=0.50, MSE = 0.0010) when applied to the held-out validation set.

**Figure 2.**
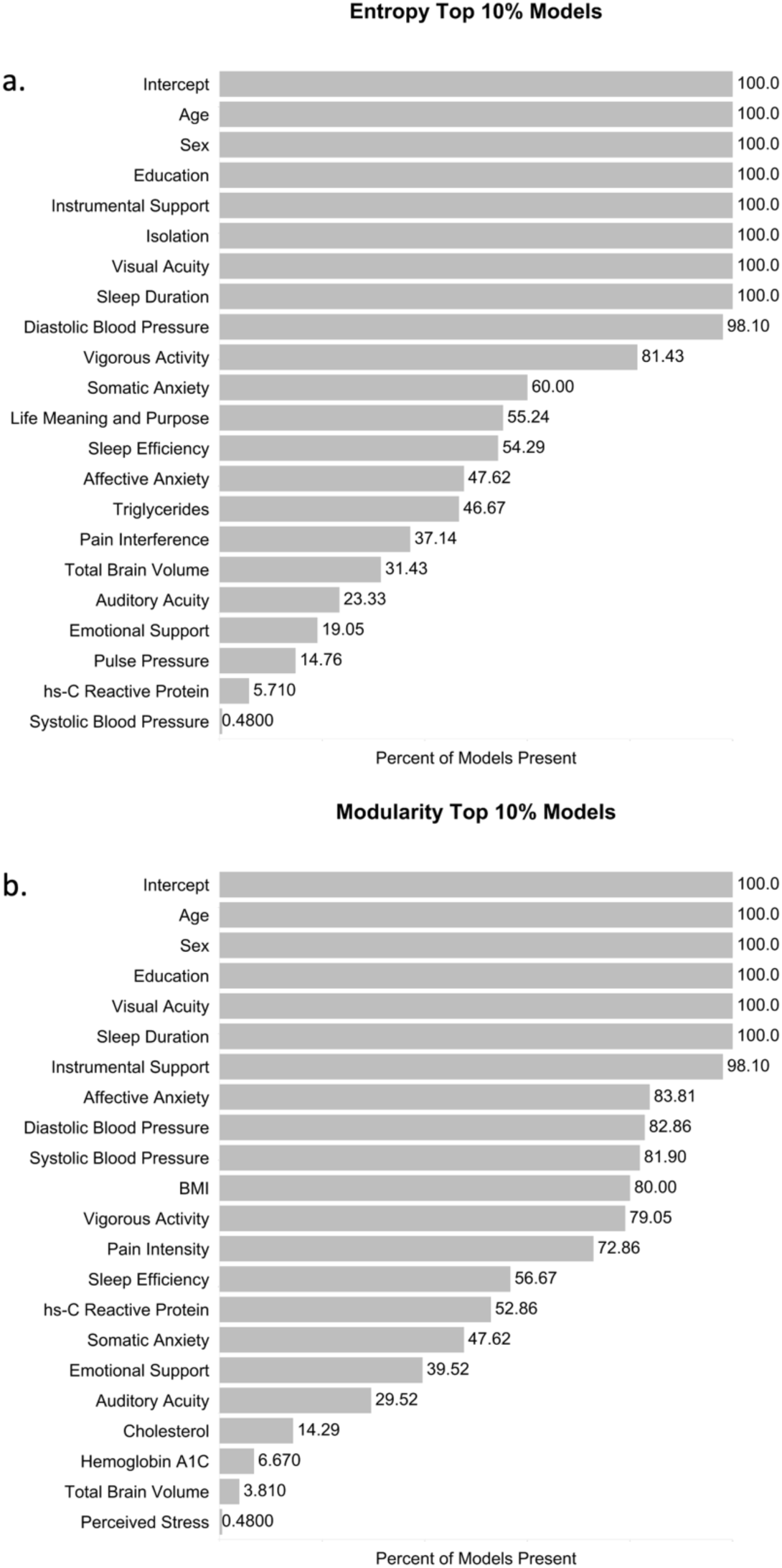
Reliability of feature selection in top 10% of models for. a) entropy and b) modularity. We demonstrate that our final model selected variables that were in the majority of the top performing models, suggesting they are reliable predictors of entropy and modularity. We also demonstrate that several features that were not selected in the final models were nonetheless selected in some of the top models, warranting further investigation in future work.

**Figure 3.**
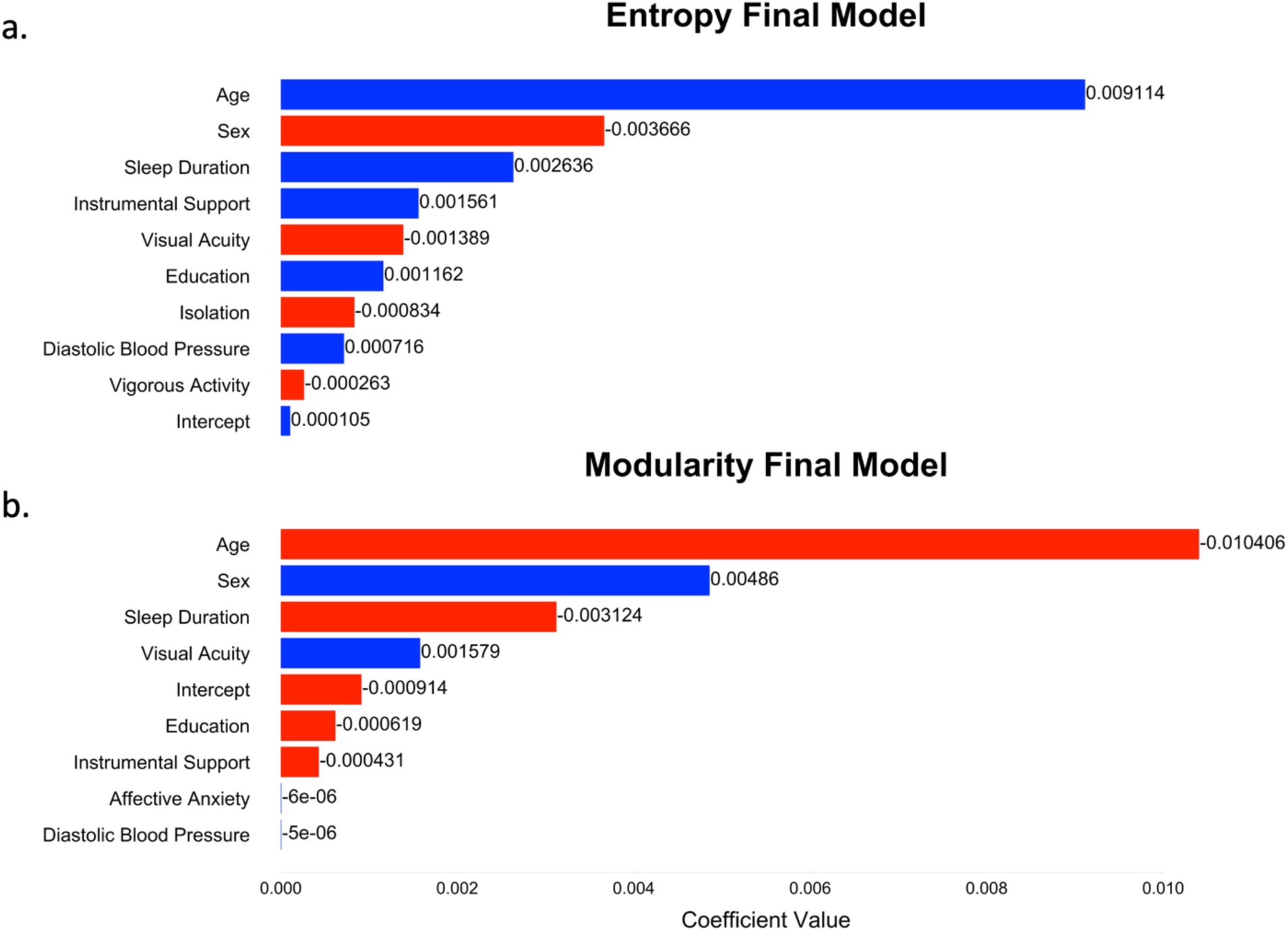
Ranking of final features selected in models for. a) entropy and b) modularity. Sex is encoded as female = 1 (male = –1).

### 3.3 Interaction Model Results

We similarly report the selected predictor variables for the top 10% of all interaction models (Figure 4). For the entropy-age interaction model, the best fitting model identified twelve final predictors (α = 1, λ = 0.0013) (Figure 5a). The top five ranked included the interaction of age and sleep hours, the interaction of age and sex, the interaction of age and education, the interaction of age and diastolic blood pressure, and instrumental support. However, despite the inclusion of twice as many predictors, model performance did not improve in the validation set (R=0.50, *r*^2^ = 0.25, MSE = 0.00054), though was slightly higher in the training set (R = 0.54; *r*^2^ = 0.30) or the. For the modularity age-interaction model, the best fitting model identified fifteen final predictors (α = 0.25, λ = 0.0076) (Figure 5b). The top five ranked included the interaction of age and sleep hours, the interaction of age and sex, the interaction of age and instrumental support, the interaction of age and education, the interaction of age and diastolic blood pressure. Model performance on the training set (R = 0.50; *r*^2^ = 0.25) and the validation set (R=0.49, *r*^2^= 0.24, MSE = 0.0010) were also similar to the simpler model.

**Figure 4.**
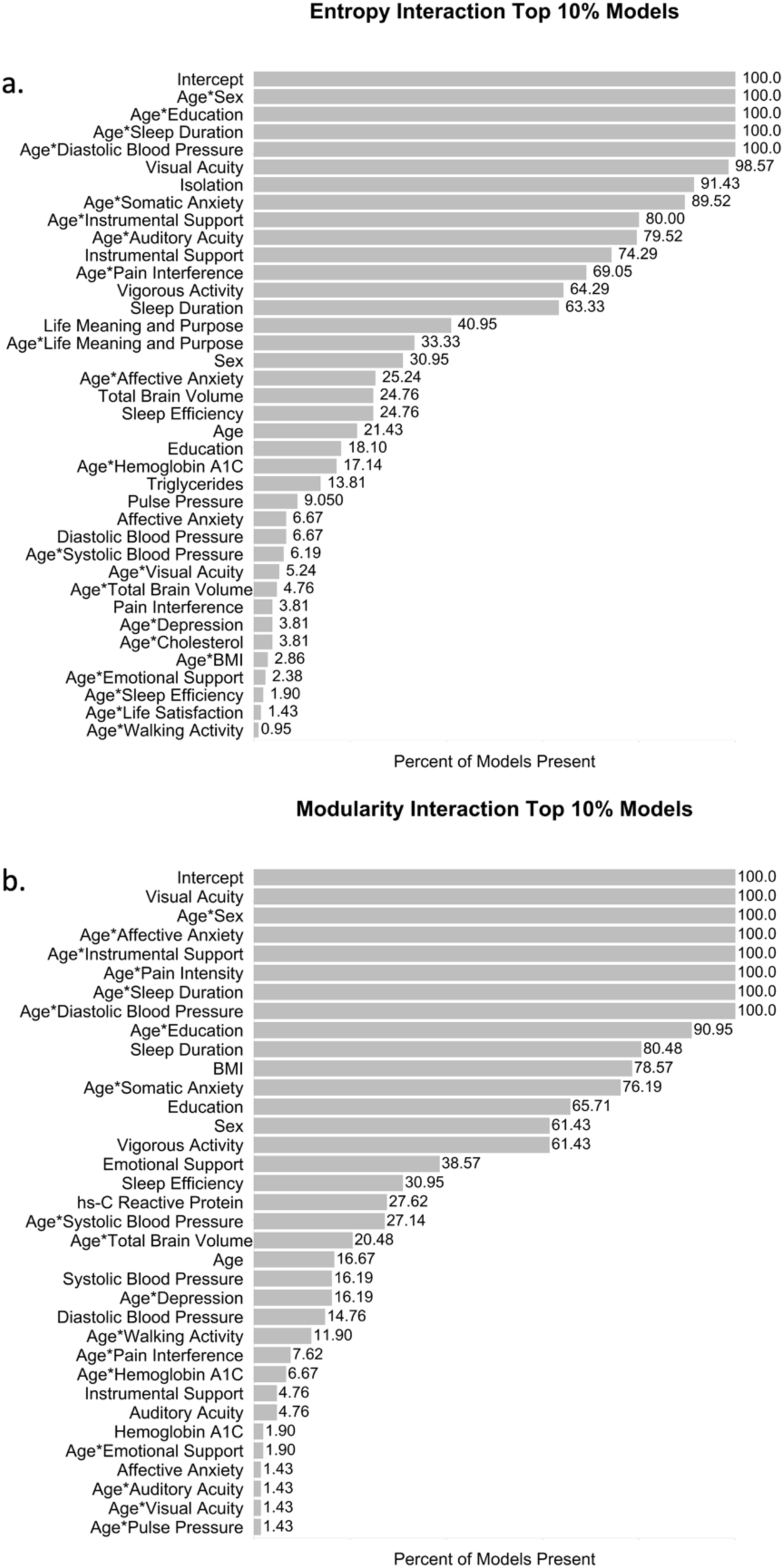
Reliability of feature selection in top 10% of interaction with age models for. a) entropy and b) modularity.

**Figure 5.**
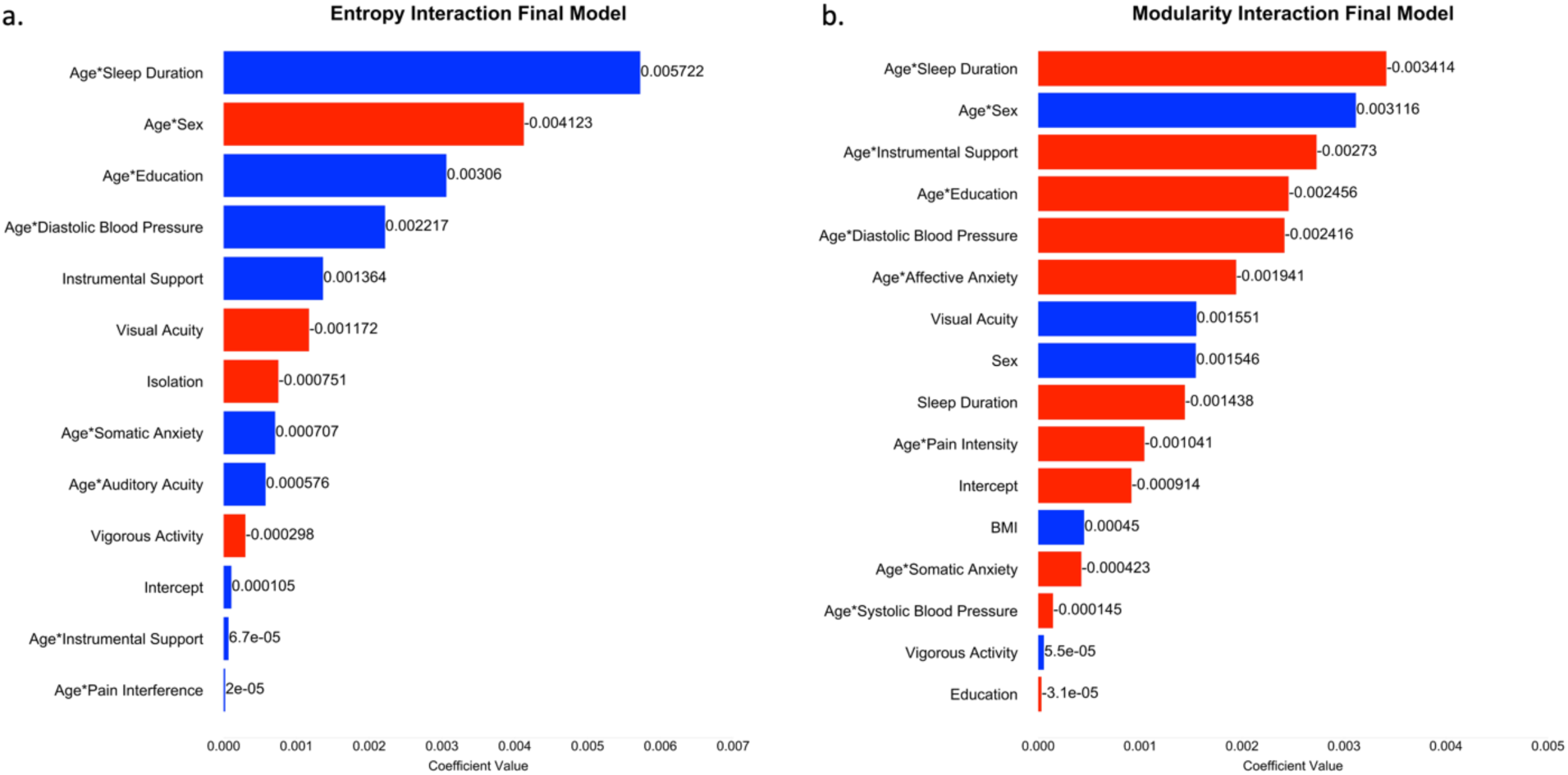
Ranking of final features selected in interaction models for a) entropy and b) modularity.

The training set performance increased only slightly, R=0.53 to R=0.54 (R=0.47 to R=0.50), for entropy (and modularity) from the simple models to the interaction models, and more importantly, performance on the validation set was identical for the entropy models and slightly worse for the modularity interaction model compared to simple model. We therefore probed the ranked interactions for predicting entropy and modularity using traditional linear regression to clarify interaction effects. We observed that only biological sex exhibited a significant interaction effect for both entropy (*B* < –0.01, t(518)= –2.40; *p* = 0.02) and modularity (*B* < 0.01, t(518)= 2.03; *p* =0.04), such that females exhibited lower entropy and higher modularity at all ages, but also demonstrated less significant age-related changes to entropy and modularity (Figure 6).

**Figure 6.**
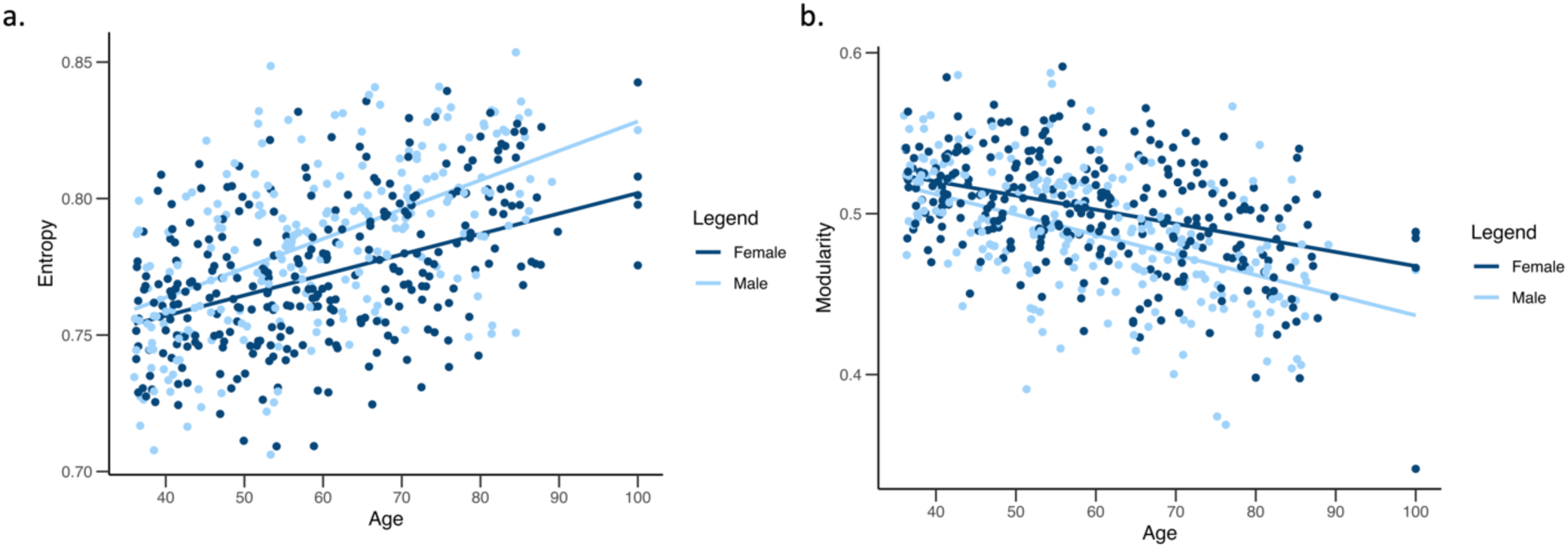
Sex differences in entropy across the adult lifespan. Interaction plots for the relationship between age and a) entropy and b) modularity by biological sex. We found that biological sex differences in entropy and modularity increase with age in this adult lifespan sample.

### 3.4 Regional Entropy

We also examined the entropy model-selected predictors at the regional brain level (Figure 7). After FDR correction to account for multiple comparisons within the 379 nodes (Benjamini & Hochberg, 1995), the predictor with the most nodes significantly correlated (*p* < 0.05) with entropy was visual acuity (Figure 7a), with 142 nodes. Of these nodes, 30.99% were observed in the temporal lobe, 23.24% in the frontal lobe, 11.97% in the cerebellum, 16.20% in the parietal lobe, 10.56% in the occipital lobe, and 7.04% in the insular cortex. Specifically, seventeen nodes in the cerebellum were significant, as well as thirteen nodes in the anterior cingulate and medial prefrontal cortex, nine in auditory association cortices, twelve in early auditory regions, fifteen in medial temporal lobe, ten in the posterior cingulate, and three in the posterior opercular regions. Notably, fourteen regions implicated in vision were significant, across early visual, dorsal and ventral visual stream, and primary visual regions. The insular and frontal opercular and orbital and polar frontal and temporo-parietal occipital junction regions were also widely implicated.

**Figure 7.**
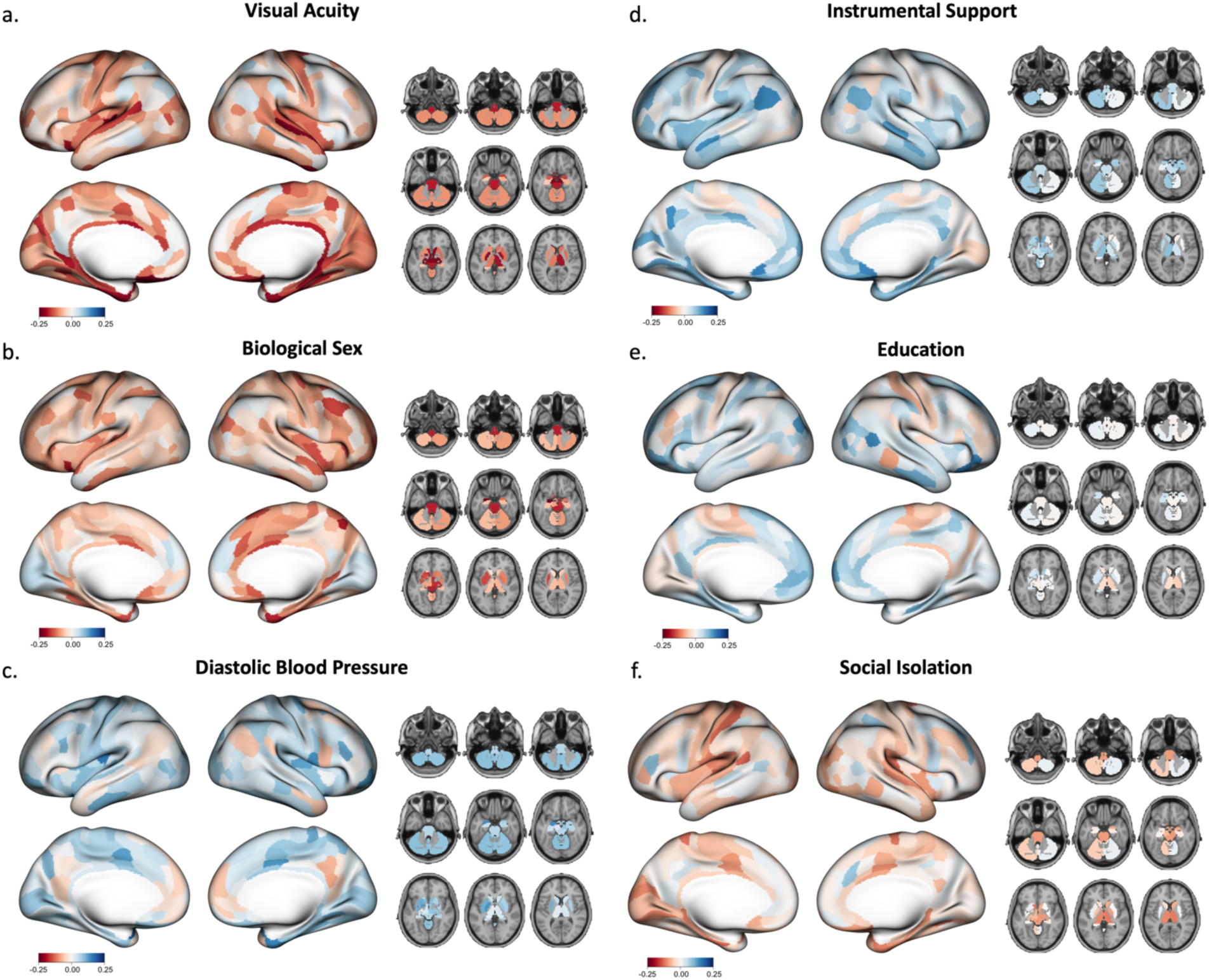
Regional entropy’s relationship with identified biopsychosocial correlates. Nodal entropy’s relationship to a) visual acuity, b) biological sex, c) diastolic blood pressure, d) instrumental support, e) education, and f) social isolation

The predictor with the second most nodes was sex (Figure 7b), with 79 significant regions. 34.18% of these regions were in the temporal lobe, 32.91% in the frontal lobe, 12.6% in the cerebellum, 11.39% in the insular cortex, and 8.86% in the parietal lobe. Cortical nodes were located primarily in the anterior cingulate and medial PFC, early auditory and auditory association areas, insular and frontal opercular regions, and medial temporal lobe. Diastolic blood pressure, the predictor with the third most significant nodes, was correlated with ten nodes: four in the frontal lobe, three in the temporal lobe, two in the cerebellum, and one in the parietal lobe (Figure 7c). Instrumental support was correlated with nine nodes, primarily in auditory regions and nodes of the anterior cingulate and medial prefrontal cortex (Figure 7d). Education was correlated with five nodes, two in superior parietal regions, one in inferior frontal cortex, one in the orbital and polar frontal region, and one in the temporo-parieto-occipital junction (Figure 7e). Social isolation was correlated with a single node in early auditory regions (Figure 7f). Despite being the third and eighth strongest predictor of entropy at the whole-brain level, respectively, neither sleep duration nor vigorous physical activity was significantly correlated with any individual nodes.

## 4. Discussion

The current study used elastic net regression to examine relationships between collinear biopsychosocial predictors and measures of functional brain network segregation and specialization. Our results provide insight into the biopsychosocial factors that may predict network segregation and nodal specialization across the lifespan. Specifically, we found that biological sex, sleep duration, instrumental support, visual acuity, education, social isolation, diastolic blood pressure, and vigorous physical activity reliably predicted whole-brain entropy across the adult lifespan. Similarly, biological sex, sleep duration, visual acuity, education and instrumental support predicted network modularity across the adult lifespan. Our results also revealed that biological sex moderated the relationship between age and network properties, such that males exhibited larger age-related changes. Previous work has hypothesized that brain networks may play a complex role in the relationship between aging and cognitive decline, both affected by pathology and therefore a mechanism of cognitive decline (Livingston et al., 2024; Reuter-Lorenz & Park, 2014), while the strength of network resilience may also serve as a buffer against that decline (Chan et al., 2021; Shankar et al., 2024). Here, we interpret our findings through this dual conceptualization and find that certain biopsychosocial factors previously linked to brain pathology predict network segregation and nodal segregation across the adult lifespan, while other predictive factors have a more complex relationship that may be more indicative of cognitive reserve.

A key advantage of elastic net regression is the ability to fit models that are generalizable and flexibly select predictors to maximize that generalizability. Our models accounted for 30% and 25% of the variance in entropy and modularity training sets, respectively, and a similar 25% and 24% variance in their respective validation sets. These results suggest our models are robust to unseen data and allow us to isolate predictors that account for the most variance in functional brain networks without compromising generalizability. Previous work examining correlates of modularity have used smaller sample sizes and traditional statistical methods (Ben Simon et al., 2017; Callow et al., 2024; Kong et al., 2020; Manza et al., 2020; Rashid et al., 2021), both of which over-inflate effect sizes and decrease generalizability to new data. Furthermore, previous work has examined only a few predictors at a time, and determined relevance through the statistical significance of relationships, biasing the identification of predictors toward those with unique, non-statistically-overlapping relationships with brain networks, and obscuring relationships between collinear variables.

Theories of brain maintenance have suggested that as individuals age, their brains accumulate damage, and that individuals who engage in health-promoting behaviors minimize the effects of age on structural and functional brain changes (Alvares Pereira et al., 2022). Here, we observed that individuals with more self-reported vigorous physical activity and lower diastolic blood pressure had lower entropy values, indicative of preserved network specialization. We found that entropy was not correlated with vigorous physical activity in any individual brain regions, indicating that the relationship between higher whole-brain averaged entropy and lower amounts of vigorous physical activity may be driven by diffuse pathological changes. Similarly, only ten brain regions were strongly correlated with diastolic blood pressure. These findings may reflect the more diffuse effects of small vessel disease on the brain (Wallin et al., 2018). Given research that higher network segregation has been positively associated with cognitive performance (Stevens et al., 2012), interventions that promote increased vigorous activity and healthy blood pressure may be important factors that promote healthy brain and cognitive aging.

We also observed that individuals with higher visual acuity had lower whole-brain-averaged entropy values and higher modularity values, indicative of preserved network segregation. Only the most recent models of dementia risk have highlighted visual acuity as an important dementia risk factor (Livingston et al., 2024). Here, we support this finding, showing that visual acuity was involved in nodal entropy values in widespread regions of the temporal lobe, including auditory cortices, and was surprisingly less commonly implicated in visual regions. This may reflect either that limiting the quality of sensory input affects brain networks, potentially leaving them more vulnerable to the effects of pathology, or that Alzheimer pathology in the temporal lobe reflects similar pathology in the retinas that affects visual acuity (Mirzaei et al., 2020), and therefore network properties are serving as a biomarker of damage accumulation.

The hypothesis that brain network properties may be an important biomarker of cognitive reserve and may explain why some individuals may demonstrate resilience to aging neuropathology has recently gained traction (Chan et al., 2021; Shankar et al., 2024; Tooley et al., 2021). We observed that individuals with long sleep duration, greater instrumental support, and less social isolation had higher entropy values, suggesting weaker average nodal specialization. Similarly, individuals with long sleep duration and more instrumental support had lower modularity values. The direction of these relationships is unexpected for instrumental support and social isolation, and we speculate that these relationships could be due to individuals with poorer overall health sleeping and experiencing less isolation as a result of their lack of independence. These factors, therefore, align with the conceptualization of modularity and entropy as biomarkers of cognitive reserve, i.e. longer sleep duration is not a cause of neural pathology but instead a compensatory behavior. An increased need for sleep has been identified as a potential behavioral symptom of dementia, neurodegeneration, and Alzheimer’s pathology accumulation (Westwood et al., 2017), perhaps due to the role in clearing these pathologies that occurs in sleep’s deepest stages (C. Wang & Holtzman, 2020), or perhaps reflective of decreased emotional or physical health leading to fatigue (Corfield et al., 2016; Williamson et al., 2005).

The 2024 Lancet Commission on Dementia prevention (Livingston et al., 2024) cites the evidence of the relationship between sleep duration and dementia risk as unclear; here we demonstrate that sleep duration consistently and strongly predicts functional network properties that likely support cognitive function. While understudied, this is consistent with recent previous work demonstrating that longer sleep duration predicted lower modularity in a sample of 135 non-demented older adults, and speaks to the need for more investigations of the relationship between sleep and functional networks (Callow et al., 2024).

We further observed that sleep duration was not significantly correlated with entropy in any single brain region despite being the strongest potentially modifiable predictor of whole-brain averaged entropy. Similarly, entropy in only a single region of the early auditory cortex was strongly correlated with social isolation, again suggesting these variables may be affected by brain networks through diffuse, rather than regional, mechanisms. Entropy in nine regions was significantly positively correlated with amount of instrumental support in regions involved in key aspects of attention, executive function, and memory (Devinsky et al., 1995; Leech & Sharp, 2014; Rodrigue & Raz, 2004), potentially reflecting that maintaining specialization in these regions is important for maintaining cognitive functions supporting independent living.

In this sample, we observed an unexpected relationship such that individuals with greater education demonstrated lower modularity and higher entropy values. Prior work has consistently characterized education as a protective factor in the development and maintenance of cognition (Chan et al., 2021; Tooley et al., 2021), though there are notable limitations to its use as a measure of cognitive reserve. Research has suggested that quality, and not quantity, of education is likely a more accurate predictor of cognitive reserve, and that quality of education has been historically very different for marginalized populations (Manly et al., 2003). This unexpected relationship may be an artifact of our highly educated, healthy sample with a large proportion of retirees, or may indicate a more complicated underlying process. Despite the unexpected direction of this relationship and its weak direct relationship, education was reliably selected in our model derivation procedure, suggesting it remains an important variable for generalizability of models predicting network properties.

Biological sex was a consistent and strong feature in all of our top models. Previous results have reported that while sex differences in modularity may be minimal in early adulthood, these differences emerge with age and increase into the early thirties and are driven by decreases to males’ modularity values (Conrin et al., 2018). A lifespan study using UK biobank data demonstrated, that while females had overall lower modularity, males demonstrated greater age-related decreases in the visual, limbic and default networks (Foo et al., 2021). In the current dataset, we found instead that females had higher whole-brain modularity and lower entropy values across the lifespan. Furthermore, with advancing age, males demonstrated more widespread age-related entropy increases across the cortex and cerebellum compared to females. Future research is needed to better understand these findings, given that women are more likely to be diagnosed with dementia and cognitive decline than their male counterparts. Emerging evidence suggests that this disparity may be influenced by differences in education and life expectancy, reduced estrogen in postmenopausal women, and experiences of discrimination (Livingston et al., 2024). The effect of these sex and gender related confounding variables on functional brain networks warrants further examination.

The interaction models containing the predictor variables and their interaction terms with age did not perform better than the simple prediction models, suggesting caution interpreting the selected interactions. Furthermore, other than the interaction of biological sex and age, the interaction terms selected in these models were nonsignificant using traditional significance testing. As with the terms selected in our simple models, these relationships warrant further, hypothesis-driven testing. However, we observed that across the entropy interaction model and the modularity interaction models, the direction of all interaction terms suggests that for older adults, functional brain networks seem to be more influenced by factors such as sleep duration, biological sex, instrumental support, education, diastolic blood pressure, anxiety, auditory acuity, physical activity, and pain than in younger adults. This provides some evidence to suggest that functional brain networks may become more malleable in response to biopsychosocial factors as age increases, which is in line with previously demonstrated large age-related changes to functional networks.

In these analyses we used machine learning to model complex relationships between functional brain network metrics and twenty-eight candidate biopsychosocial predictors. The advantage of elastic net regression is that it can identify and rank predictors of outcome variables even when those predictors are collinear. For example, in our raw correlations between variables, we observed strong to moderate relationships between network properties and features such as auditory acuity, BMI, Hemoglobin A1C, systolic blood pressure, and pulse pressure.

However, none of these were selected in our top performing models. We also did not observe relationships between mental health variables and entropy, and though the best fitting model for modularity selected affective anxiety as a predictor, this was not reliably selected in all models and had an effect size smaller than the intercept. These findings are somewhat surprising, given findings that depression, in particular, has an effect on brain networks (Gong & He, 2015b). It is therefore important to acknowledge the limitation of self-report data in the assessment of mental health diagnoses and symptoms of depression and anxiety and that these findings may differ in research where mental health diagnoses and symptoms are assessed by qualified clinicians.

Additionally, some of our selected variables, such as education, demonstrated weak relationships to our outcome variables but were nonetheless reliably selected in the top performing models. This suggests that these variables impart important predictive information even if they are weakly directly related. Elastic net regression is also a penalized regression technique, meaning it is less prone to model overfitting and more likely to produce generalizable findings. Indeed, our models accounted for a similar proportion of the variance in entropy and modularity in our validation set as they did in our training set.

The prioritization of generalizability and prediction, however, does not necessarily equate to selection for variables based on their effect size. While effect sizes may be small in the current sample, at a population level these factors may account for a not insignificant opportunity to bolster dementia prevention efforts. Additionally, the inclusion of age interaction terms did not increase model performance despite the statistical significance of at least one of these terms, suggesting that our predictor to sample size ratio was not large enough to address this question definitively. Larger and more diverse samples may reveal additional variables that may be equally or more influential in determining network properties and their change across the lifespan. Future studies are also needed in additional datasets to confirm the generalizability of the results presented in this study. Finally, the low signal-to-noise ratio of fMRI data means that we cannot make strong claims about the differences between our models predicting entropy and modularity, especially given that these metrics are themselves highly correlated.

Despite these limitations, the current study identifies several key features predictive of brain network function across the lifespan in addition to age: sex, sleep duration, instrumental support, visual acuity, education, social isolation, diastolic blood pressure, and vigorous physical activity. This finding elucidates the factors that may contribute most notably to cognitive reserve in aging.

## Data Statement

All data are from a publicly available dataset, the Human Connectome Project Lifespan 2.0 Release.

## CRediT authorship contribution statement

**Anita Shankar:** Conceptualization, Formal Analysis, Data Curation, Writing-Original Draft, Writing-Review & Editing, Visualization. **Madhura Phansikar**: Validation, Data Curation, Writing-Review & Editing. **Nathan McPherson:** Validation, Writing-Review & Editing. **Adam X. Gorka:** Conceptualization, Writing-Review & Editing. **Baldwin Way:** Writing-Review & Editing. **Richard Betzel:** Conceptualization, Writing-Review & Editing. **Ruchika S. Prakash:** Conceptualization, Funding Acquisition, Writing-Review & Editing, Supervision

## Data Availability

The study used publicly available data from NDA.org the Human Connectome Project Lifespan Release. All data produced in the present study are available upon reasonable request to the authors.

## Supplemental Methods

This publicly available dataset aimed to collect data on ‘typically’ aging adults, including those with common health conditions such as hypertension, but excluding individuals with more serious conditions, including those with neurological disorders, macular degeneration, or significant cognitive deficits for their age (Bookheimer et al., 2019). Participants underwent a blood draw, saliva collection, breathalyzer, and urine sample collection to obtain genotyping, metabolic, lipid, and hormonal data after a recommended 8 hour fast (Bookheimer et al., 2019). Participants also completed a physical examination and were assessed for vascular health factors. Finally, participants completed a series of comprehensive self-report questionnaires assessing aspects of biopsychosocial health factors, including the Perceived Stress Scale (Cohen et al., 1983), Pittsburgh Sleep Quality Index (Buysse et al., 1989), International Physical Activity Questionnaire (Craig et al., 2003), PROMIS Social Isolation (Hahn et al., 2014) and the NIH Toolbox Emotion Battery (Salsman et al., 2013), from which we selected subtests that tapped domains with prior associations with dementia: Life Satisfaction, Emotional Support, Affective Fear, Somatic Arousal, and Life Meaning and Purpose. We also included tests of sensation and sensory acuity from the NIH Toolbox Sensation Battery: Pain Intensity and Pain Interference (Cook et al., 2013), the Visual Acuity Test (Varma et al., 2013), and the Words-In-Noise Audition Test (Zecker et al., 2013). Participants also completed two, 45-minute MRI sessions, which included the collection of structural images (T1w, T2w) and functional images during rest, a Go/No-Go attentional task, a Visual-Motor Processing Speed task, and a Face-name pairing associative memory task. Imaging at all study sites used a Siemens 3 Tesla Prisma scanner and a 32-channel head coil that enabled multiband data acquisition (Harms et al., 2018). From this dataset, we included 28 predictors including age, years of education, and sex as demographic variables. Participants were excluded from analysis if they had missing data on any of the fMRI or predictor variables. Twelve participants were missing at least one task fMRI scan and two were missing intracranial volume data. Eleven were excluded for missing at least partial demographic data. Ninety additional individuals were excluded for missing blood draw data. Seven additional individuals were excluded for missing physical exam data, and seventy-eight individuals were excluded for missing in the self-report data on at least one of the examined measures.

Blood draw measures included blood triglycerides, cholesterol, high sensitivity C-Reactive Protein (hsCRP), and Hemoglobin A1c (HbA1c). Cardiovascular health measures included body mass index, systolic blood pressure, diastolic blood pressure, and pulse pressure. From the self-report questionnaires, we included the total raw scores from the Perceived Stress Scale, sleep efficiency and duration from the Pittsburgh Sleep Quality Index, and frequency of walking, moderate and vigorous physical activity from the International Physical Activity Questionnaire. From the NIH and PROMIS toolboxes, we included the total scores for measures of depression, affective anxiety, somatic anxiety, instrumental support, emotional support, social isolation, life satisfaction, life meaning and purpose, pain intensity, pain interference, auditory acuity, and visual acuity. We also include total brain volume as a control variable given the observed sex differences in brain volume.

## Supplemental Results

For models predicting entropy and modularity without regressing out motion, we observed slightly higher model fit accuracy to our primary results but observed some changes to selected predictor variables. For entropy, the best fitting model from the derivation procedure identified sixteen final predictor variables ( α= 0.75, l= 0.001), ranked in order of importance to model predictive power as: age, sex, BMI, hours of sleep, diastolic blood pressure, visual acuity, instrumental support, education, social isolation, somatic fear, hsCRP, total brain volume, sleep efficiency, A1CRS, pain interference, and vigorous physical activity. This model’s performance yielded an r-squared of 0.29 (R= 0.54, MSE = 0.00058) when applied to the 156 individuals in the held-out validation set. For modularity, the best fitting model identified twenty-one final predictor variables ( α = 0.20, l = 0.0048), ranked as: age, sex, visual acuity, hours of sleep, systolic blood pressure, instrumental support, diastolic blood pressure, hsCRP, pain intensity, auditory acuity, sleep efficiency, affective anxiety, emotional support, vigorous physical activity, BMI, education, somatic anxiety, triglycerides, cholesterol, depression, and A1CRS. The model when applied to the 156 individuals in the held-out validation set yielded an r-squared of 0.29 (R=0.54, MSE =0.00103) when applied to the held-out validation set.

